# Immediate, remote smoking cessation intervention in participants undergoing a targeted lung health check: QuLIT2 a randomised controlled trial

**DOI:** 10.1101/2022.04.26.22274257

**Authors:** Parris J Williams, Keir EJ Philip, Navjot Kaur Gill, Deirdre Flannery, Sara Buttery, Emily C Bartlett, Anand Devaraj, Samuel V Kemp, Jamie Addis, Jane Derbyshire, Michelle Chen, Katie Morris, Anthony A. Laverty, Nicholas S Hopkinson

## Abstract

**Background:** Lung cancer screening programs provide an opportunity to support smokers to quit, but the most appropriate model for delivery remains to be determined. Immediate face to face smoking cessation support for people undergoing screening can increase quit rates, but it is not known whether remote delivery of immediate smoking cessation counselling and pharmacotherapy in this context is also effective.

**Materials and Methods:** In a single-blind randomised controlled trial, smokers aged 55-75 years attending a Targeted Lung Health Check (TLHC) were allocated by day of attendance to receive either immediate telephone smoking cessation support (TSI) (starting immediately and lasting for 6 weeks) with appropriate pharmacotherapy, or usual care (very brief advice to quit and signposting to smoking cessation services) (UC). The primary outcome was self-reported 7-day point prevalence smoking abstinence at three months. Differences between groups were assessed using logistic regression.

**Results:** 315 current smokers taking part in the screening programme, mean (SD) age 63(5.4) years, 48% female, were randomised to telephone smoking cessation (n=152) or usual care (n=163). The two groups were well-matched at baseline. Self-reported quit rates were higher in the intervention arm, 21.1% vs 8.9% (odds ratio [OR]: 2.83, 95% CI 1.44-5.61, p=0.002). Controlling for participant demographics, baseline smoking characteristics or the discovery of abnormalities on low dose CT scanning did not modify the effect of the intervention.

**Conclusion:** Immediate provision of an intensive telephone-based smoking cessation intervention, delivered within a targeted lung screening context, is associated with increased smoking abstinence at three months.

**Trial registration:** This study is registered online: ISRCTN12455871.

**Take home message:** Providing immediate, telephone smoking cessation support with pharmacotherapy to smokers enrolled in a TLHC program increases quit rates in this population by almost a third.

## Introduction

Targeted screening programmes using low dose computed tomography (LDCT) have been proposed as a solution to reduce the impact of lung cancer, by diagnosing it at an earlier, potentially curable stage. Large randomised controlled trials have demonstrated that this approach can reduce mortality from lung cancer by between 20-60% (1, 2) and LDCT screening is now recommended by the US Preventative Services Taskforce (3). In 2019, NHS England launched targeted lung health check (TLHC) pilot projects at various sites across the UK to investigate the feasibility and effectiveness of LDCT screening within the UK (4), more recently the National Screening Committee (NSC) began a public consultation regarding a national roll out of TLHC across the UK(5).

Tobacco smoking is among the largest causes of morbidity and mortality and smoking cessation is thus a key aspect of the prevention and treatment both of respiratory disease and many conditions occurring beyond the lungs (6). Lung cancer screening eligibility criteria targets “high risk smokers”, who differ in certain ways from the general smoking population being older, often with multiple co-morbidities, a longer smoking history and greater tobacco dependence (1, 7). LDCT screening trials have demonstrated higher quit rates in intervention than control arms and the screening process can thus be considered to be a “teachable moment” for smoking cessation (8, 9). Making the best use of this is crucial, and the provision of evidence-based smoking cessation within screening programmes has been advocated (10). The effectiveness of different approaches remains a key question for research, to establish which specific approaches should be employed to maximise the value and impact of the lung health check (11-14).

In the initial phase of the Quit Smoking Lung Intervention Trial (QuLIT-1) we demonstrated a significant increase in 3 month quit rate (29.9% vs 11%), for TLHC participants randomised to receive immediate face to face cessation support and pharmacotherapy, compared to usual care (UC). The latter consisted of very brief advice to quit (VBA) and signposting to smoking cessation support(15). Because of the COVID-19 pandemic, face to face support was suspended in March 2020 and the study was modified to investigate, in a discrete second phase (QuLIT-2), whether remote delivery of the smoking cessation intervention was also effective compared to usual care.

## MATERIALS AND METHODS

### Study design and Participants

QuLIT-2 was a single-blind, randomised, controlled trial comparing the effectiveness of an immediate, intensive telephone smoking cessation intervention (TSI) compared to Very Brief Advice (VBA) and signposting delivered in the context of a TLHC. People living in the London boroughs of Sutton, Hillingdon, and Hammersmith and Fulham, aged 55-75, with a recorded history of smoking, were invited for a TLHC assessment as described previously (15). All current smokers who took part in the TLHC were included in the study (current smokers were defined as any person self-reporting smoking tobacco including cigarettes, pipes and cigars at the time of the TLHC).

### The Targeted Lung Health Check Setting

The Healthy Lung Project is an investigational lung cancer screening pilot delivered by the Royal Brompton Hospital, supported by RM Partners, the West London Cancer Alliance, and NHS funded through the National Cancer Transformation Fund. Due to the COVID-19 pandemic, in 2021 all initial pre-scan TLHC appointments were changed to a remote telephone delivery model. They included an in-depth discussion of participants’ current or historical smoking behaviour, medical and occupational history, and familial cancer history. If participants were deemed high risk of lung cancer according to the Prostate, Lung, Colorectal and Ovarian (PLCO) or Liverpool Lung Project (LLP) screening risk models (16, 17) they were invited for a LDCT.

### Ethics

The study was approved by the South Central – Oxford C Research Ethics Committee and the Health Research Authority (Ref:18/SC/0236). The requirement for individual consent was waived by the ethics committee, as obtaining this would itself have been an intervention and influenced outcomes in the control group. The initial QuLIT pilot study (QuLIT-1) and the amended study reported here (QuLIT-2) were registered prospectively online (ISRCTN12455871).

### Randomization

Participants in the trial attended their TLHC appointment between 1^st^ April and 31^st^ June 2021. Half the days during that period were allocated by random number generation as Telephone Smoking Intervention days and half as Usual Care. Appointments for the TLHC were allocated by an administrator who was unaware of which study arm days had been allocated to.

### Interventions

#### Telephone Smoking Cessation

Participants in the TSI group received a telephone call from the smoking cessation practitioner after their initial TLHC appointment. To capitalise on this “teachable moment” the smoking cessation practitioners attempted to call the participants the same day they received their TLHC. If they were unable to get through, they would call the participant the next day. Participants were offered six sessions of telephone behavioural counselling support, in addition to pharmacotherapy (varenicline or nicotine replacement therapies).

Counselling sessions were based on the National Centre for Smoking Cessation and Training (NCSCT) and KickIT (18, 19) programmes. The initial session included a discussion of participants’ smoking history, involving an assessment of dependence, information about available pharmacotherapy and information about the program. The session would finish with a summary of the pharmacotherapy chosen and a commitment from the participant to engage with the program. Before a Varenicline presciption was made the nurses discussed the patients’ medical history with a medical doctor (KEJP or NSH) who would then prescribe the medication or suggest an alternative. All pharmacotherapies were prescribed by the trial team and despatched via the Hospital’s pharmacy. We sent the chosen pharmacotherapy to participants directly by mail immediately after the first session, typically arriving within 48 hours.

The second session included preparing the participant to set a quit date and ensuring they knew how to use/ take their chosen pharmacotherapy, and a discussion around typical withdrawal symptoms. Sessions 3-6 were delivered after the set quit date and would include support with withdrawal symptoms, pharmacotherapy reviews, commitment to the “not a puff” rule for participants who had quit and further behavioural support for participants who relapsed or were unable to commit to their quit date. All sessions were delivered over the phone by two specialist research nurses who had undergone the National Centre for Smoking Cessation Training (NCSCT) and KickIT training programmes (18, 19).

#### Usual Care

Those attending on usual care days received VBA to quit (“Stopping smoking is the most important thing that you can do to improve your health now and reduce the risk of health problems in the future.”) as outlined by the NCSCT (20). They were directed to the London Stop Smoking Portal https://london.stopsmokingportal.com/ which provides information to smokers about how to engage with local stop smoking services, as well as a Quitline service. Participants living in the London borough of Sutton were advised to get in touch with their GP for stop smoking support as there was no specialist smoking cessation support available in this borough. VBA to quit and signposting were delivered by the specialist respiratory nurses who administered the TLHC clinics and occurred at the end of their appointment.

#### Follow up

Three months following the TLHC appointment, participants were called by a researcher (KEJP or PJW) who was blind to study allocation. The content of the call was structured on a set of short-predefined questions. This included the primary outcome measure which was self-reported 7-day point prevalence smoking abstinence, with a successful quit attempt defined as no smoking or other tobacco product use within the last 7 days. Data relating to secondary outcome measures was also collected, including quit attempts made, pharmacotherapy used, and experiences of the service as described previously. If participants did not pick up on the first call, two more calls were made at different times of the day. If the participant did not pick up on the third call, a voicemail was left requesting a call back. In the event that the participant did not call back within the week (or had no voicemail facility) they were classed as lost to follow up.

#### Statistical analysis

The primary outcome was self-reported 7-day point prevalence abstinence from smoking, 3 months post randomization compared between groups. The sample size was calculated using the findings of two studies; the EAGLES trial (21) which found a 38% quit rate in the pharmacology arm and the UK Lung Cancer Screening Study (8), which found a 14% quit rate in the arm undergoing CT screening. Based on these rates, a superiority study (1:1 randomisation) with 90% power at a 5% significance level would require 136 participants. To improve the power of exploratory analyses comparing different sub-groups (e.g. those with or without positive CT results), we intended to recruit as many participants from the clinical screening program as possible. Simple logistic regression analysis (unadjusted) was used to assess primary and secondary endpoints. We ran 2 models in the adjusted logistic regression analysis, model 1 adjusted for sex, age and CT result and model 2 adjusted for sex, age, and baseline demographics (age, sex, smoking characteristics). As a sensitivity analysis, we assumed all individuals lost to follow up were still smoking. Analysis was based on intention to treat, and a p value of <0.05 was taken as statistically significant. All data were analysed using SPSS V27.

## RESULTS

Baseline participant characteristics at the time of enrolment were well matched (Table 1). A total of 315 smokers underwent a TLHC during the study period and were enrolled onto the study, 163 attended on days allocated to UC and 152 on TSI days, figure 2 represents the flow of patients through the trial.

**Table 1.** Baseline Demographics

| **Characteristics** | **Usual Care (n=163)** | **Intervention (n= 152)** |
| --- | --- | --- |
| Age, mean years (SD) | 63.0 (5.4) | 61.3 (4.8) |
| **Sex** |  |  |
| Female, n (%) | 84 (51.5) | 68 (44.7) |
| Male, n (%) | 79 (48.5) | 84 (55.3) |
| **Ethnicity, n (%)** |  |  |
| White | 138 (84.6) | 131 (86) |
| Black | 9 (5.6) | 6 (4.0) |
| Asian | 12 (7.3) | 9 (6.0) |
| Other | 3 (1.9) | 5 (3.3) |
| Prefer not to say | 1 (0.6) | 1 (0.7) |
| **Education, n (%)** |  |  |
| < GCSE Level | 64 (39.3) | 59 (38.9) |
| GCSE Level | 63 (38.7) | 49 (32.2) |
| A Level | 23 (14.2) | 17 (11.1) |
| Some University | 2 (1.3) | 7 (4.6) |
| Degree + Postgrad | 11 (6.5) | 20 (17.2) |
| **IMD quintile n (%)** |  |  |
| 1 (Most deprived) | 9 (5.5) | 15 (9.9) |
| 2 | 19 (11.7) | 19 (12.5) |
| 3 | 40 (24.5) | 26 (17.1) |
| 4 | 65 (39.9) | 52 (34.2) |
| 5 (Least Deprived) | 30 (18.4) | 40 (26.3) |
| **Baseline Smoking Characteristics** |  |  |
| Age started smoking, mean (SD) years | 18.1 (6.3) | 18.0 (5.9) |
| Average number of cigarettes per day, mean (SD) | 12.6 (7.9) | 12.5 (7.7) |
| Number of smoking years, mean (SD) | 42.5 (9.5) | 40.9 (9.0) |
| **CT Result** |  |  |
| No Scan needed, n (%) | 59 (36.2) | 52 (34.2) |
| CT negative, n (%) | 55 (33.7) | 60 (39.4) |
| CT positive, n (%) | 49 (30.1) | 40 (26.4) |

**Ethnicity groupings as recommended by the Prostate, Lung Colorectal and Ovarian Screening Trial (PLCO).**
**Index of multiple deprivation quantile (IMD), quantile 1= most deprived areas in England quantile 5= least deprived areas in England (IMD) (22).**

### Engagement with smoking cessation

Of the 152 participants attending on days randomised to TSI, 80 (52%) declined to engage with smoking cessation, explicitly asking not to be contacted by our cessation nurses. Of the remaining 72 participants in the TSI arm, 57/73 (78%) 72(79%) were enrolled onto the full cessation program and 16/72 (21%) dropped out after the initial session. Reasons given for dropping out were being unable to commit to the “not a puff” rule, or not being ready to commit to the program.

### Outcomes

Three month follow up data were available for 227/315 (72%) participants (UC: 115/163 (70%), TSI: 112/152 (73%). Quit rates were higher in the intervention arm; 21.1% vs 8.9% (OR: 2.83, 95% CI 1.44-5.60, p=0.002) (Table 2). The number of quit attempts reported, including successful and unsuccessful attempts was also higher in the TSI group (57/152 37.5% vs 36/163 22.0%), (OR: 2.11, 95% CI 1.29-3.47 p=0.003), (Table 2, Figure 1). We explored if receiving the CT scan itself influenced quit rates among the study arms. Within the UC arm receiving a CT scan did not influence quit rates (OR, 0.68, 95%CI 0.20-2.28, p= 0.53), 4/59 (6.7%) quit rate among UC participants who did not receive a CT scan and 10/104 (9.5%) among UC participants who recived a CT scan. This was similar within the TSI study arm, 14/52 (26.9%) quit rate among TSI participants who did not receive a CT compared to 18/100 (18%) quit rate among TSI participants who recived a CT, (OR, 1.67, 95%CI 0.75-3.72, p= 0.23).

**Table 2.** Smoking cessation and quit attempts at 3-months

|  | **Telephone quit**<br>**smoking**<br>**intervention**<br>**(n=152)** | **Usual care**<br>**(n=163)** | **Odds Ratio (95%**<br>**CI)** | **P Value** |
| --- | --- | --- | --- | --- |
| Seven-day smoking abstinence | Yes 32 (21%)<br>No 120 (79%) | Yes 14 (8.9%)<br>No 149 (91.1%) | 2.83 (1.44-5.61) | 0.002 |
| Individuals reporting a quit attempt | Yes 57 (37.5%)<br>No 95 (62.5%) | Yes 36 (22.0%)<br>No 127 (78.0%) | 2.11 (1.29-3.47) | 0.003 |
| Pharmacotherapy Aids used to<br>support quit attempts |  |  |  |  |
| • Varenicline | 16 (28.0%) | 1 (2.7%) | 13.65 (1.72-108.24) | 0.01 |
| • Single item NRT | 12 (21.0%) | 2 (5.5%) | 4.53 (0.95- 21.61) | 0.05 |
| • Dual item NRT | 21 (37.0%) | 3 (3.8%) | 6.41 (1.75-23.51) | 0.005 |
| • E-cigarette | 8 (14.0%) | 11 (30.5%) | 0.37 (0.13-1.04) | 0.05 |
| • Nil | 0 (0%) | 19 (57.5%) | 0.06 (0.0-0.27) | 0.004 |

***NRT nicotine replacement therapy (NRT)******CI, Confidence interval***

**Figure 1.**
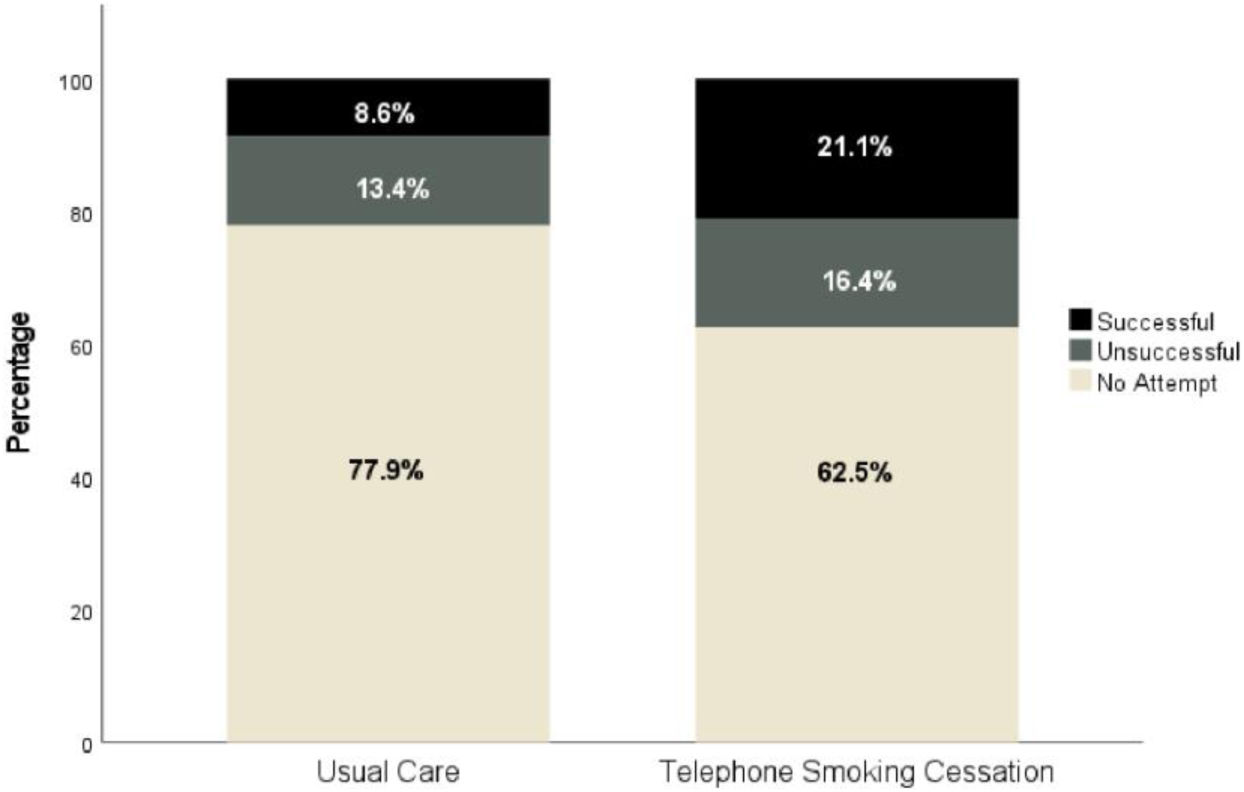
Percentages of quit attempts made per study arm, including no quit attempt, successful quit attempts and unsuccessful quit attempts made.

**Figure 2.**
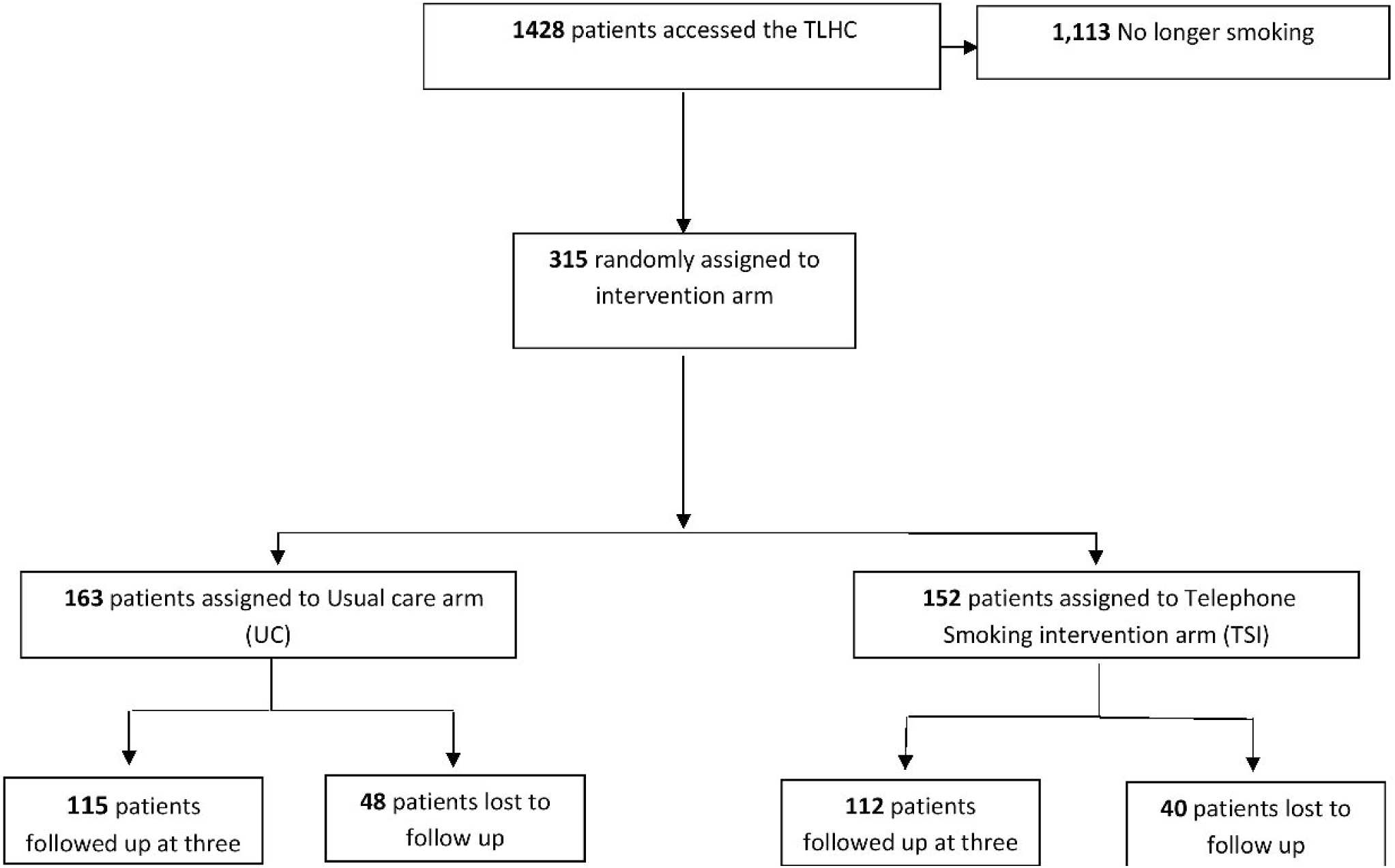
Flow diagram to represent the flow of patients through the QuLIT2 trial

The use of all types of pharmacotherapies (varenicline, single item NRT and dual item NRT) during quit attempts was more common in the TSI group (OR: 20.90, 95% CI 6.98-62.55, p=<0.001) (Table 2). By contrast, participants in the UC group were more likely to report use of e-cigarettes to aid quit attempt (TSI: 14.0% vs UC: 30.5%). Of note, in the UC arm only 3/36 participants attempting to quit accessed behavioural support via their local service (2 quit successfully) and 1/36 used the NHS stop smoking smartphone app.

Results from the two logistic regression models are displayed in Table 3, data from participants who did not receive a CT scan were excluded from model 2. Person demographics (sex, age), CT scan result (positive, negative) and smoking characterises (average number of cigarettes per day, age started smoking and number of years smoking) had no effect on quit rates at three months within our cohort.

**Table 3.** Logistic Regression Models of covariates associated with smoking abstinence at 3 months

|  | **Model 1: Sex, age, CT result**<br>**Odds ratio (95% CI)** | **Model 2: Sex, age, baseline**<br>**smoking characteristics Odds**<br>**ratio (95% CI)** |
| --- | --- | --- |
| **Intervention** | **2.80 (1.42-5.53)** | **2.62 (1.32-5.24)** |
| Sex (men vs. women) | 1.00 (0.581-1.44) | 1.00 (0.57-1.43) |
| Age (continuous) | 1.00 (0.90-0.98) | 1.01 (0.90-1.01) |
| CT Negative | 1.46 (0.77-2.72) | - |
| CT Positive | .990 (0.50-1.61) | - |
| Average Number of cigarettes<br>per day | - | 0.995 (0.97-1.04) |
| Age started Smoking | - | 1.02 (0.95-1.07) |
| Number of years Smoking | - | .977 (0.94-1.03) |

## DISCUSSION

The main finding of this randomised controlled trial is that the provision of smoking cessation support including counselling support and pharmacotherapy, delivered by telephone immediately following attendance at a targeted lung health check, significantly increased 7-day point prevalence smoking abstinence at 3 months, compared to usual care of very brief advice to quit and signposting to smoking cessation services.

### Significance of findings

Our results support the hypothesis that the immediate provision of remote, intensive smoking cessation support for high-risk people who smoke undergoing TLHC, is more effective (21.1% vs 8.9%) for increasing quit rates within this cohort than usual care. These findings extend those from our previous trial QuLIT-1, which found immediate face-to-face support can increase quit rates compared to usual care (29.2% vs 11%) in the context of TLHC (15). Comparing the two trials, the quit rates in the usual care groups in both studies were numerically similar whereas quit rates in the face-to-face intervention were higher. The number of quit attempts was also higher in the intervention group (TSI: 37.5% vs UC: 22.0%) and similar between both QuLIT-1 and QuLIT-2 interventions (QuLIT-1: 35.3%). However, the two approaches were not directly compared, so future studies will be required to confirm which approach is generally more effective.

Several trials, including our initial QuLIT-1 study have investigated the impact of smoking cessation support within lung cancer screening pathways (11, 12, 14, 15, 23) The 21.1% three-month quit rate observed in the QuLIT-2 intervention arm is comparable with other studies where quit rates ranged from 10-25%. Interestingly, the two studies which reported lower interventional quit rates than our current study conducted their cessation remotely (telephone and online) and did not offer any pharmacotherapy (11, 23).

The results of previous studies have been mixed, limited by factors including statistical power and the nature, intensity, duration, and immediacy of intervention. Marshall et al (14), randomized 55 smokers to either one session of smoking cessation counselling delivered face to face or printed self-help materials and direction to a Quitline on the day of screening. They found no significant difference in quit rates between their intervention (14.3%) and control arms (16.4%). This may be attributable to the low intensity interventions in both arms, and the small size of the pilot study.

In a study of internet self-help smoking cessation materials, delivered on the day of screening compared to standard information leaflet and Quitline in 171 smokers who underwent low-dose fast spiral chest CT screening, Clark et al. found significantly higher quit attempts at 1 year follow up (68% vs 48%)(11). This is mirrored somewhat by our current study with higher quit rates among the TSI arm compared to UC (37.5% vs 22.0%), of note the cessation intervention used by Clark et al, is drastically different to ours, more investigation is needed to explore quit attempts within this context.

Other studies have also investigated which factors might be associated with successful smoking cessation within TLHC settings. Tremblay et al (13) found that those who reported a cancer history, smoked at work, and consumed alcohol once a week or more were more likely to be still smoking at 12 months (13). In the NELSON screening trial, smokers with higher educational attainment and higher motivation to quit were more likely to have quit at 2 year follow up (24). By contrast, we found no associations between participant characteristics and successful smoking abstinence, although this may be due to a lack of statistical power. The effect of receiving a CT scan vs, not having a CT as part of a TLHC did not significantly alter quit rates within our population (CT: 13.7% vs No CT: 16.2%). These data do support the offer of smoking cessation support to everyone taking part in targeted lung health screening, not only those whose risk score means that they qualify for low dose CT scanning.

Pharmacological cessation aids were accessed by people trying to quit in both study arms, however participants in the TSI group were around 20 times more likely to use pharmacological aids including NRT and varenicline. This is unsurprising as the combination of pharmacological aids and behavioural support were the main components of our intervention. Yet the pharmacotherapy usage within the UC arm was remarkably low only 6/36 (16%) participants who attempted to quit used pharmacotherapy and only 3 participants in the UC arm accessed cessation support from their local services despite advice and signposting to do so. E-cigarette usage to support quit attempts was significantly higher in the UC arm of our study (TSI: 14.0% vs UC: 30.5%), indicating that in the absence of organised support and pharmacotherapy, smokers are more likely to adopt this approach. There are data to support the effectiveness of e-cigarettes in the context of lung cancer screening, with significantly higher 3 month abstinence with both nicotine e-cigarettes(25.4%) and nicotine free e-cigarettes (23.4%) compared to control (10.34) (25). These findings coupled with the recent announcement from the UK’s Medicines and Healthcare Products Regulatory Agency (MHRA) that it will support the medicinal licensing of e-cigarettes for smoking cessation,(26) suggest that providers of cessation interventions within a screening context should be prepared to support people using this form of nicotine replacement.

Several studies have investigated remotely delivered smoking cessation support, and a 2013 Cochrane review investigating the effectiveness of telephone support concluded that proactive telephone counselling may be useful for increasing access to cessation support particularly if the calls are an adjunct to other self-help materials and provided more than once (27). The Alberta lung screening trial reported no significant difference in 12 month quit rates between 345 smokers randomised to seven sessions of telephone behavioural support, compared to Quitline and information leaflets (12.3% vs 14.0%)(13). Zelidit et al, compared the effectiveness of two smoking cessation behavioural support calls conducted after participants had received their CT results, to national Quitline number (control group) and reported substantially higher quit rates at 30 days in the intervention (19%) compared with control (7%) (23). Finally, a smaller study of 93 participants, which randomised participants to either six sessions of telephone behavioural support or standard information leaflet and Quitline smartphone app, demonstrated an increased quit rate with telephone behavioural support (17.4% vs 4.3%) (12). Both studies which reported significantly higher quit rates in their intervention arms, and, as well as our own, delivered cessation support within the first few days of participants’ TLHC appointment or results, whereas the Alberta screening trial intervention was delivered around 16 days post screening. It should be noted that the current study is the only one out of those discussed that made pharmacotherapy treatment available as part of the intervention.

Our results have the potential to inform future service delivery of TLHC programs through optimisation of smoking cessation support delivery in this population. Both QuLIT-1 and QuLIT-2 suggest improved quit rates with early intervention delivery, either face-to-face or remotely, compared to usual care. Although face-to-face approaches might be more effective for those able to attend in person, remotely delivered interventions could expand access in general and are of particular interest during the COVID-19 pandemic where limitations have been placed on face-to-face activities. It is likely that a combination of delivery options will be most appropriate for a population with varying needs and preferences.

### Strengths and limitations

We conducted the study in the context of a clinical lung cancer service, without deviation from the standard pathway or change in patient experience, apart from the provision of the smoking cessation support on TSI days, which increases the generalisability of the findings. The inclusion of all current smokers enrolled in the healthy lung project, regardless of their current readiness or motivation to quit, allows us to assess the impact of the approach within the whole screened population, not just those immediately motivated to stop smoking.

Certain limitations and considerations should be discussed. Firstly, we used self-reported 7-day point smoking prevalence as our primary endpoint rather than biochemically confirmed quit rates to keep study costs and participant burden low. Although point prevalence self-reported data is a known valid abstinence measure within clinical trials (28), and often used in other cessation screening trials as an outcome measure (8, 12, 13). The use of exhaled CO monitoring would increase the rigor of the study outcome. Thirdly, the loss to follow up observed (28%) within our population was slightly higher than anticipated, though similar to other studies (11, 25). Importantly, sensitivity analysis, taking the cautious assumption that all those lost to follow up continued to smoke, did not alter the study findings.

## Conclusion

Immediate, intensive telephone-based smoking cessation support with pharmacotherapy, delivered within a TLHC setting increased three month quit rate. This suggests that this approach is appropriate and effective for this population and that access to specialist smoking cessation support should be embedded within the delivery of lung cancer screening.

## Data Availability

All data produced in the present study are available upon reasonable request to the authors

## DATA SHARING

Anonymised research data will be shared with third parties via request to the senior author (NSH).

## AUTHOR STATEMENT

NSH, EB, AS, EB, SK, JA, JD, MC, KM designed the study, NKG and DF delivered the smoking cessation intervention, supported by PJW. PJW and KEJP conducted follow up calls, PJW, KEJP and AL conducted data analysis. PJW produced the first draft to which all authors contributed. All authors have reviewed and approved the final version. NSH is the guarantor.

## TRANSPARENCY DECLARATION

Nicholas Hopkinson, the manuscript’s guarantor affirms that the manuscript is an honest, accurate, and transparent account of the study being reported; that no important aspects of the study have been omitted; and that any discrepancies from the study as planned (and, if relevant, registered) have been explained

